# Rapid Whole Genome Characterization of High-Risk Pathogens Using Long-Read Sequencing to Identify Potential Healthcare Transmission

**DOI:** 10.1101/2024.08.19.24312266

**Authors:** Chin-Ting Wu, William C. Shropshire, Micah M Bhatti, Sherry Cantu, Israel K Glover, Selvalakshmi Selvaraj Anand, Xiaojun Liu, Awdhesh Kalia, Todd J. Treangen, Roy F Chemaly, Amy Spallone, Samuel Shelburne

## Abstract

**Objective:** Routine use of whole genome sequencing (WGS) has been shown to help identify transmission of pathogens causing healthcare-associated infections (HAIs). However, the current gold standard of short-read, Illumina-based WGS is labor and time-intensive. In light of recent improvements in long-read Oxford Nanopore Technologies (ONT) sequencing, we sought to establish a low resource utilization approach capable of providing accurate WGS-based comparisons of HAI pathogens within a time frame allowing for infection prevention and control (IPC) interventions.

**Methods:** WGS was prospectively performed on antimicrobial-resistant pathogens at increased risk of potential healthcare transmission using the ONT MinION sequencer with R10.4.1 flow cells and Dorado basecalling algorithm. Potential transmission was assessed via Ridom SeqSphere+ for core genome multilocus sequence typing and MINTyper for reference-based core genome single nucleotide polymorphisms using previously published cut-off values. The accuracy of our ONT pipeline was determined relative to Illumina-based WGS data generated from the same genomic DNA sample.

**Results:** Over a six-month period, 242 bacterial isolates from 216 patients were sequenced by a single operator. Compared to the Illumina gold-standard data, our ONT pipeline achieved a Q score of 60 for assembled genomes, even with a coverage rate of as low as 40X. The mean time from initiating DNA extraction to complete genetic analysis was 2 days (IQR 2-3.25 days). We identified five potential transmission clusters comprising 21 isolates (8.7% of all sequenced strains). Combining ONT WGS data with epidemiological data, >70% (15/21) of the isolates originated from patients with potential healthcare transmission links.

**Conclusions:** Via a stand-alone ONT pipeline, we detected potentially transmitted HAI pathogens rapidly and accurately, aligning closely with epidemiological data. Our low-resource method has the potential to assist in the efficient detection and deployment of preventative measures against HAI transmission.

## Introduction

Healthcare-associated infections (HAIs) cause tens of thousands of deaths and cost around $3 and $27 billion annually in England and the U.S. respectively [1]. Infection prevention and control (IPC) teams are critical to mitigating transmission of HAI pathogens, but typical IPC methodologies heavily rely on the intuition of infection control professionals, are time-consuming, and can either over- or under-identify outbreaks [2]. As whole genome sequencing (WGS) becomes more affordable and feasible, routine WGS has proven effective in detecting clusters of pathogens not meeting typical IPC criteria for potential HAI transmission. For example, Sundermann *et al.* found that over 10% of isolates were part of genetically related clusters, and only 44% of the isolates identified to be genetically related would be classified as healthcare-associated transmissions using standard National Healthcare Safety Network (NHSN) criteria [3, 4]. Similarly, Australian investigators used WGS to discover that over 30% of patients acquired MDR pathogens from the hospital [5]. Other research has demonstrated the effectiveness of WGS in distinguishing between methicillin-resistant *Staphylococcus aureus* (MRSA) outbreaks and pseudo-outbreaks [6, 7]. These WGS-based efforts highlight the importance of timely identification of HAI transmission in order to facilitate outbreak control by IPC teams. A major barrier to real-time HAI analysis using WGS is the reliance on highly accurate Illumina short-read sequencing, which requires extensive preparation and batch processing, particularly for institutions that do not have large sequencing facilities [8].

Oxford Nanopore Technologies (ONT) long-read sequencing is an alternative to Illumina short-read sequencing and generally requires minimal sample preparation, can readily be adapted to varying numbers of strains being sequenced, and provides read lengths that facilitate complete genome assemblies including plasmids [9]. Whereas ONT previously had unacceptably high error rates for assessing bacterial genetic relatedness, advancements like improved V14 chemistries, double-sensor R10 nanopores, and enhanced consensus basecalling models have significantly increased accuracy [10]. These improvements suggest that ONT sequencing could be a viable standalone real-time WGS HAI analysis option. In this study, we aimed to establish an ONT-only sequencing pipeline capable of rapidly and accurately producing WGS data to classify the genetic relatedness among potentially transmitted antimicrobial-resistant (AMR) pathogens in a tertiary care cancer hospital.

## Methods

### Study population

The study took place at the University of Texas MD Anderson Cancer Center (MDACC), a 760-bed tertiary care facility in Houston TX, USA. The study timeframe was from August 2023 to March 2024 and was approved by the MDACC quality improvement institutional review board. To optimize the chances of identifying transmitted MDR pathogens, we studied organisms previously identified at high-risk of HAI transmission, namely MRSA, vancomycin resistant *Enterococcus faecium* (VREfm), and carbapenem-resistant forms of *Enterobacterales*, *Acinetobacter baumannii* and *Pseudomonas aeruginosa* [4, 11]. Screening of the electronic health record was performed twice weekly to identify bacteria of interest with final inclusion being limited to those isolated from patients hospitalized for ≥ 48 hours at the time of infection onset or in patients with extensive recent contact (≤ 30 days) with the MDACC healthcare system (i.e. admitted to the hospital or undergoing an outpatient procedure).

### Genomic DNA extraction, long-read sequencing, and data analysis

Genomic DNA (gDNA) was extracted directly from plates obtained from the MDACC clinical microbiology laboratory using the GenElute™ Bacterial Genomic DNA Kit. Long-read libraries were prepared using the Rapid Barcoding Kit 96 V14 and were sequenced on the MinION device using R10.4.1 flow cells following manufacturer instructions. All pod5 reads were basecalled using Dorado v0.5.1 in super high accuracy mode (dna_r10.4.1_e8.2_400bps_sup@v4.3.0) with a minimum quality score filter of 8 to produce FASTQ files. Dorado v0.5.1 was also used to demultiplex and remove adapters from the sequencing results (GitHub: https://github.com/nanoporetech/dorado). Long-read assemblies were generated using an in-house Flyest package nanopore consensus polishing pipeline (GitHub: https://github.com/wshropshire/flyest). AMR gene presence was assessed using AMRFinderPlus v3.11.14 [12].

### Accuracy assessment of stand-alone ONT sequencing

Illumina sequencing was performed on 55 (22%) samples using the NextSeq500 platform at the MDACC core sequencing facility with a target sequencing depth of ∼100x. Trimmed Illumina short reads were aligned to the consensus sequence assembled from ONT long reads, and variants indicating potential errors in ONT sequencing or assembly were identified using Snippy v4.6.0 with default variant calling parameters (https://github.com/tseemann/snippy). Q score was calculated as: Qscore = -10 * log10 (total number of variants identified by Snippy / genome length assembled by Flyer). We utilized Rasusa v0.8.0 (GitHub: https://github.com/mbhall88/rasusa) to subsample our ONT FASTQ files to coverages of 100x, 80x, 60x, 40x, and 20x for the 35 strains with at least 100x ONT coverage. The subsampled data were used to generate assembled genomes, and the Illumina data were used to identify potential errors as described above.

### Measures of genetic relatedness

Sequence types (ST) and clonal complexes (CC) were determined by PubMLST databases. To identify strains with sufficient genetic similarity to indicate potential transmission, we conducted a two-step screening with cutoff values for different species derived from previously published studies (Table 1) [13–16]. First, SeqSphere+ software (Ridom SeqSphere+ version 9.0.10) was used for core genome multilocus sequence typing (cgMLST) as the primary screening method. The FASTA file from the ONT assembly was utilized, requiring at least 95% of cgMLST target genes for subsequent analysis. For strains that met the cgMLST cut-off, reference-based single nucleotide polymorphism (SNP) calling was performed using MINTyper version 1.1.0 directly on the FASTQ files generated from Dorado demultiplexing [17]. For strains where cgMLST schema are currently not available in SeqSphere+ (e.g. *Enterobacter cloacae*), genetic relatedness was exclusively assessed using MINTyper with cut-offs derived from previously published data [18].

**Table 1:** Cutoff values for two-step screening.

|  | Primary screening |  | Secondary analysis |
| --- | --- | --- | --- |
|  | Number of Seqsphere+<br>cgMLST target genes | cgMLST cutoff<br>(allele) | core-genome SNP<br>cutoff (SNP) |
| *S. aureus* | 1861 | <24 | < 24 |
| *E. faecium* | 1423 | < 20 | < 20 |
| *E. coli* | 2513 | < 15 | < 15 |
| *K. pneumoniae* | 2358 | < 15 | < 15 |
| *P. aeruginosa* | 3867 | < 20 | < 40 |

### Combination of genetic and epidemiologic data

For strains that met our potential transmission genetic cut-off values, transmission likelihood was classified based on previously published definitions with probable transmission assigned to patients who stayed on the same ward with at least 24 hours of overlap, possible transmission being for patients who stayed in the same ward within 60 days without overlap, and unlikely transmission when neither of the above criteria were met [5]. Figure 1 illustrates the entire workflow of our ONT sequencing process.

**Figure 1.**
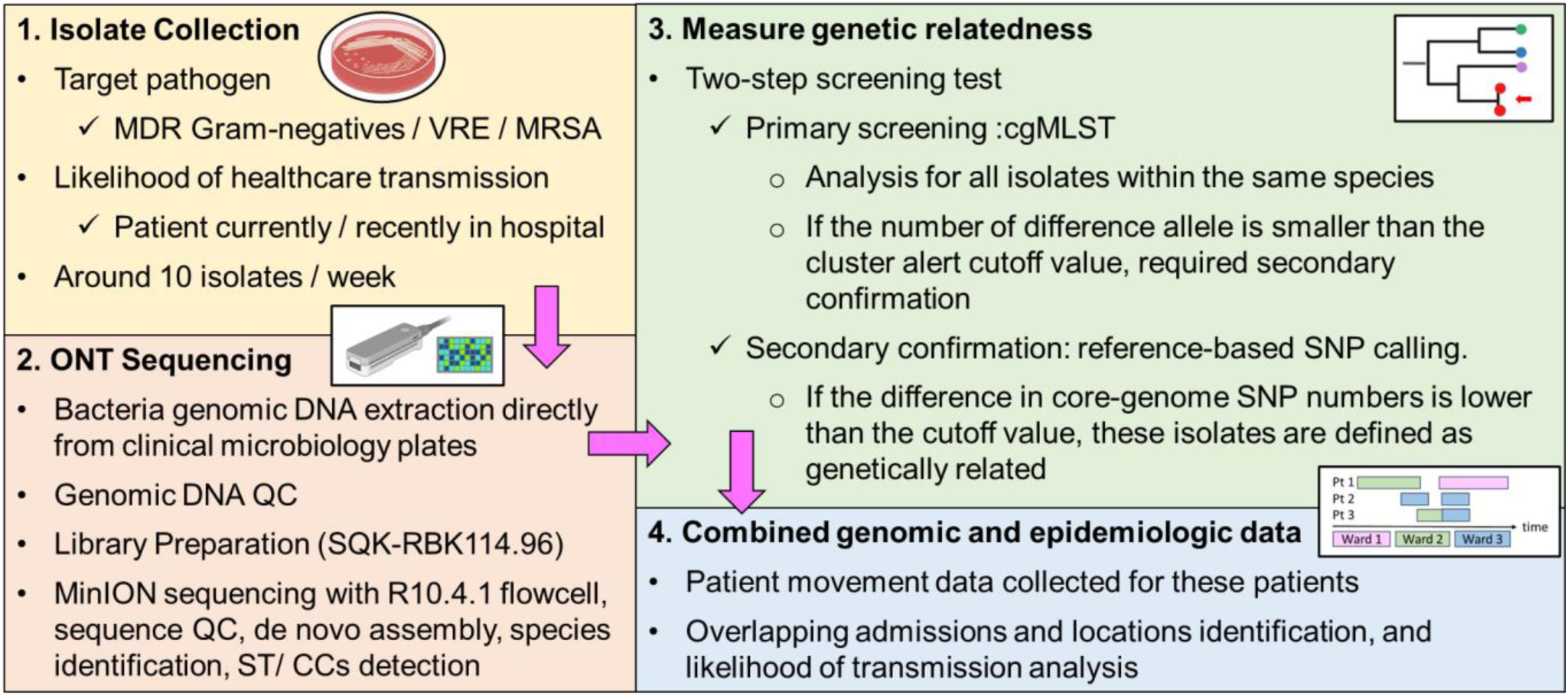
Workflow of stand-alone real-time ONT sequencing pipeline.

## Results

### Standalone ONT sequencing data accuracy

From August 2023 to March 2024, we performed ONT sequencing on 242 unique clinical isolates from 216 patients with a species breakdown of 83 MRSA (33%), 41 VREfm (17%), 37 *P. aeruginosa* (15%), 31 *E. coli* (13%), 26 *K. pneumoniae* (11%), and 24 other species (10%) (Table S1). Once we had performed ONT sequencing of at least ten isolates of the five top species, we performed parallel Illumina sequencing on the same genomic DNA. We mapped the Illumina reads to the genomes assembled with ONT data to determine the number of SNPs/ insertion-deletion (INDELs) (i.e. errors) in the ONT assemblies. The median number of SNPs was 1, interquartile range 1 to 3 (IQR): 0-2 and the median number of (INDELs) was 3 (IQR: 1-5) (Figure 2A). The average Q score for the ONT-assembled genomes was 60.7 (IQR: 56.9-64.5). We observed that 87% of SNPs were transitions, with 42% being T to C and 33% being A to G (Figure 2B). The outlier in Figure 2A, a ST584 *K. pneumoniae* sample, exhibited 60 SNPs, with 31 being A to G and 29 being T to C. The Integrative Genomics Viewer of an example SNP is shown in Figure 2C. We conclude that relative to the Illumina gold standard, the ONT sequencing was highly accurate although still with a low level of A to G and T to C transitions likely due to poor modeling of unique methylation motifs [19].

**Figure 2:**
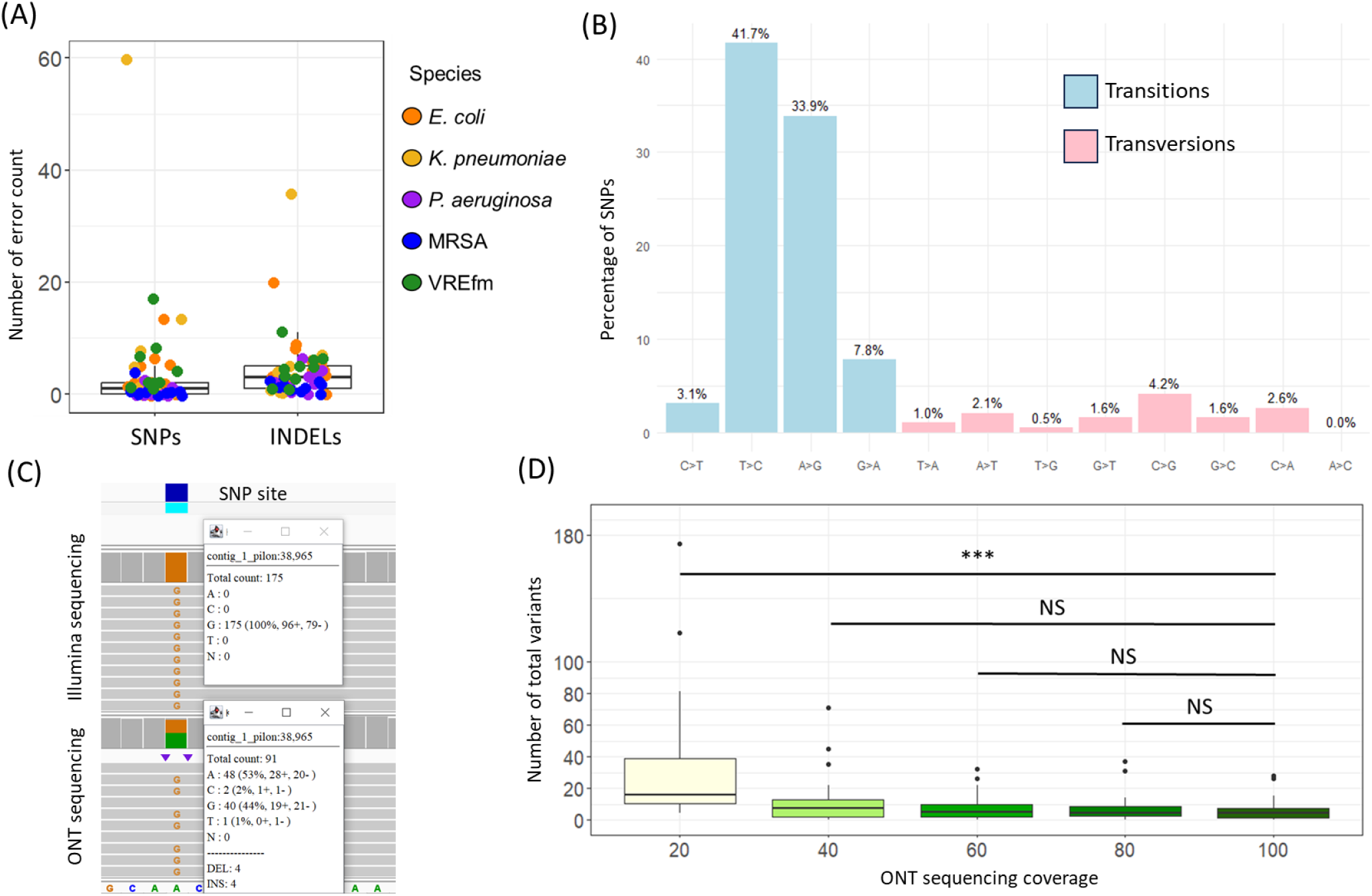
Standalone ONT sequencing data accuracy validation. (A) Number of variants (i.e. errors) in standalone ONT pipeline vs Illumina data (n = 55 strains). Errors are classified into single nucleotide polymorphisms (SNPs, left) and insertion/deletions (INDELs, right). Colors indicate strain species shown in legend. (B) SNPs were classified as transitions (light-blue) or transversions (pink) with exact genetic variation shown on X-axis. Numbers refer to the percentage of total SNPs made up by each genetic variation. (C) Example of SNP error site. Upper part of panel shows Illumina reads mapped to ONT assembly where 100% Illumina reads map to referent (i.e., Adenine, A) as an alternative allele (i.e., Guanine, G) and lower panel indicates ONT alignment with approximately a 50% A/G mapping. (D) Impact of sequencing coverage on error rates with ONT sequencing depth used to generate ONT assembly shown on X-axis and total number of errors shown on Y-axis (median and IQR are shown). *** *P*-value < 0.001. NS = no statistical difference for indicated Pairwise Wilcoxon Rank-Sum test results.

Higher sequencing coverage and coverage depth generally improves assembly accuracy by reducing the likelihood of misassembly or missing sequences [20]. However, targeting lower coverage depth per sample allows for sequencing more samples per flow cell, which can be cost-effective [20]. Thus, we next aimed to optimize sequencing coverage to balance cost-effectiveness and accuracy. We used Rasusa to subsample the ONT files to achieve 100X, 80X, 60X, 40X, and 20X coverage. We found that the total number of variants was significant higher at 20X coverage relative to 100X coverage (P < 0.001 by Kruskal-Wallis test followed by pairwise Wilcoxon rank-sum test), but no significant differences in total variants were observed for coverages ≥ 40X (Figure 2D).

### Execution of our standalone ONT sequencing workflow

Using our ONT sequencing workflow (Figure 1), over a 26-week study period, we screened 494 positive AMR culture reports. Of these, 246 met our inclusion criteria with three strains excluded due to discrepancies between species ID in the clinical microbiology laboratory and by sequencing analysis whereas one strain was excluded due to sequencing failure leaving a total of 242 isolates for analysis. The most common isolate source was blood (36%, 88/242), followed by tissue/wound/body fluid (24%, 58/242), urinary tract (20%, 48/242), and respiratory tract (18%, 44/242). On average, 10 strains were sequenced per week. The fastest time from initiating gDNA extraction to completing data analysis was one day, with a mean duration of two days (IQR: 2-3.25 days). Detailed timelines of the workflow and sequencing quality control data are provided in Figure S1 and Table S1.

### Overview of major sequence types amongst sequenced pathogens

Phylogenetic trees for each of the five major species are shown in Figures S2-6. Among the 31 *E. coli* isolates, half belonged to ST131 (n=16, 51.6%) followed by ST167 (n=4, 13%) and ST361 (n=2, 7%). The 26 *K. pneumoniae* isolates displayed diverse STs with ST258 being the most common (n=4, 15%), followed by ST45 (n=3, 12%) and ST147 (n=3, 12%). For 37 *P. aeruginosa* strains, over 19% (n=7) were identified as ST633, while other isolates had unique sequence types. ST117 (25 of 41 total isolates, 61%) dominated the VREfm population followed by ST80 (n=6, 15%). Most MRSA isolates (n = 83) were from CC8 (n=38, 46%), CC5 (n=30, 36%), and CC30 (n=8, 10%). A heat map of major genes mediating acquired β-lactam resistance is shown in Figure S7. Except for the large numbers of *P. aeruginosa* ST633 strains, our AMR pathogen epidemiology generally aligned with predominant STs/CCs reported globally [21–25].

### Characterization of potential transmission using genetic relatedness assessment

Using our two-step screening process, we identified 21 isolates (8.7%) meeting the criteria for possible transmission, forming five genetically related clusters (Figure 3). Among these, three clusters were from ST117 VREfm and one from ST80 VREfm, with 46% of VREfm strains (19/41) meeting the genetic relatedness cut-off for possible transmission. Another cluster involved two ST45 *K. pneumoniae* isolates. Table S2 details the detected clusters. To visualize potential healthcare transmissions, we used patient-ward-movement timelines to integrate genomic and epidemiological data. Figure 4 shows the largest ST117 VREfm genetic-related cluster, comprising 12 unique patients. Within this cluster, seven patients (A, C, D, E, H, J, and L) shared overlapping stays in ward 1 and ward 4 for at least one day, indicating a probable transmission. Two patients (B and I) were on ward 2 at different times but within 60 days, suggesting a possible transmission. However, three patients (F, G, and K) did not have overlapping admissions or locations. Potential epidemiological links were present in >70% (15/21) of isolates that met our genetically related cut-off values.

**Figure 3:**
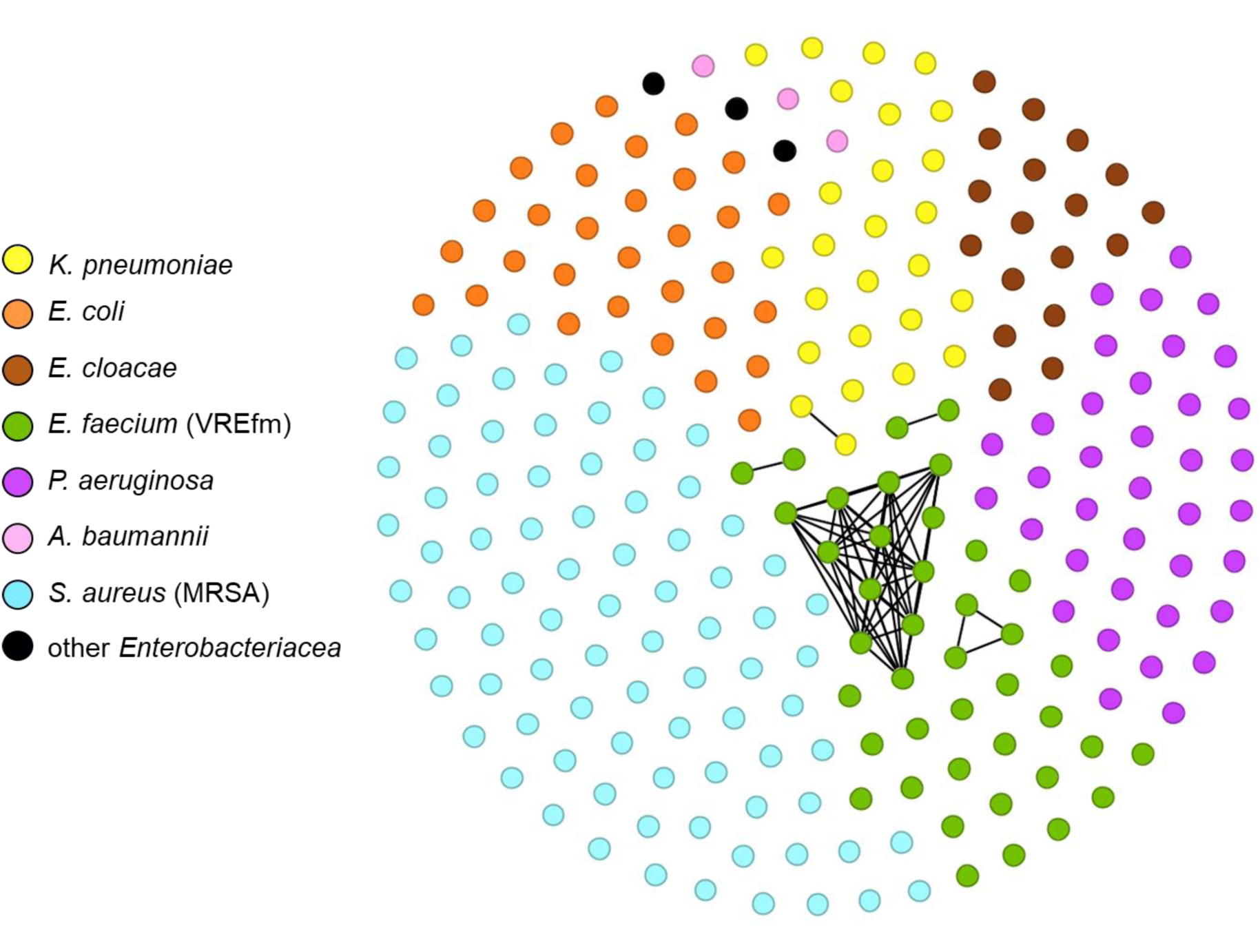
Genetic-related cluster network found in our standalone ONT sequence pipeline. Each dot represents one sequenced isolate, color-coded by bacterial species. The network plot was visualized using Gephi. Dots on the outer circle without lines indicate isolates above the cutoff values, while dots in the inner circle, connected by lines, indicate strains below the cutoff values, suggesting possible transmission.

**Figure 4.**
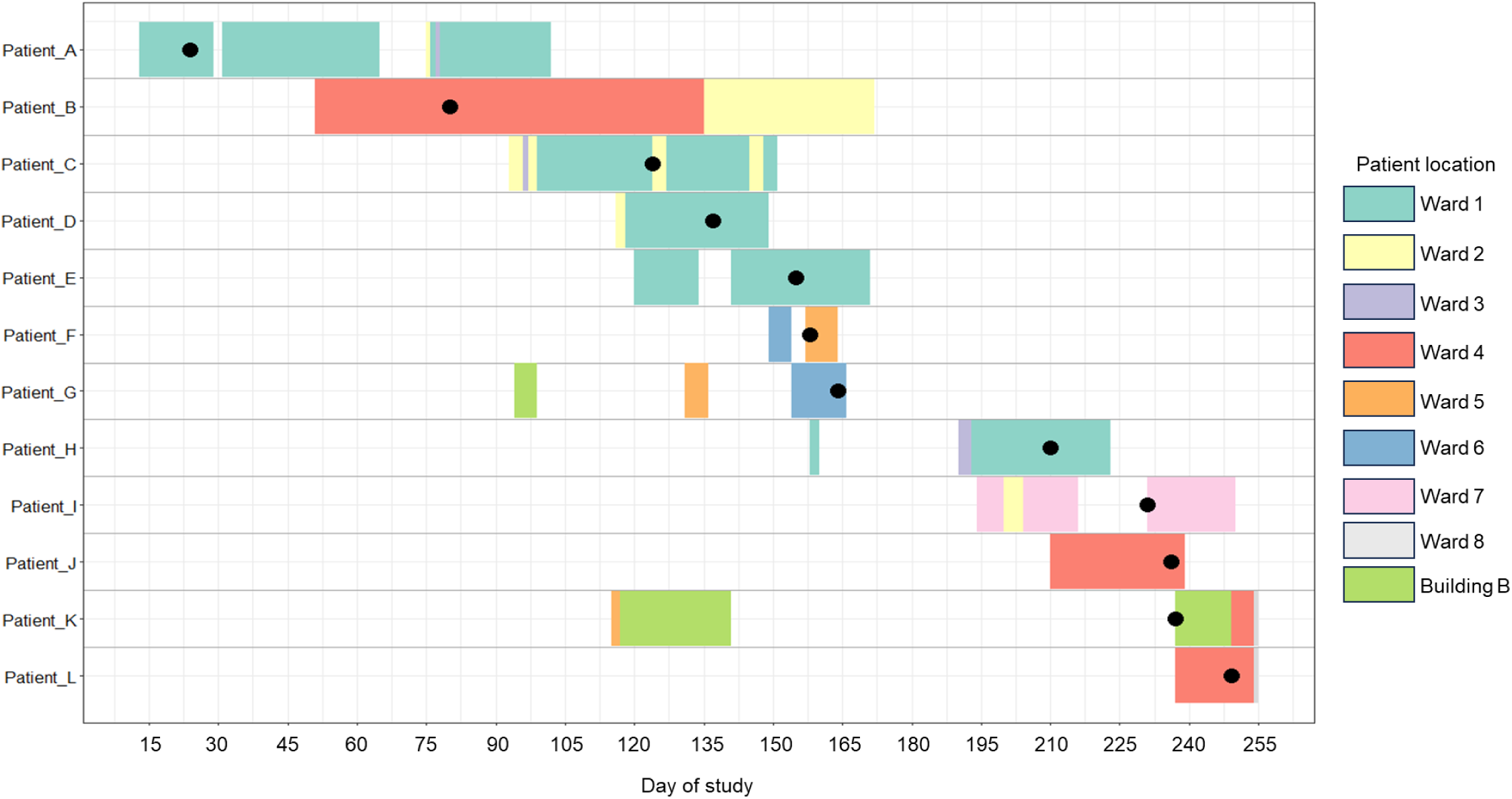
Patient-Ward-Movement-Timelines for VREfm Cluster IV. Twelve unique patients in cluster IV are listed on the left. Day of study is shown on the x-axis. Colored boxes in the figure indicate the time-ward-movement timeline for the 12 patients with the individual colors as shown in the legend on the right. Black circle dots represent sample collection date.

### Use of ONT data to assess potential plasmid transmission

Given the power of long-read ONT sequencing to assess plasmid composition [26], we also sought to determine whether our ONT sequencing pipeline could assess the possibility of plasmid transfer among different bacterial strains/species. We focused on plasmids containing *bla*_NDM_ genes, which encode New Delhi metallo-β-lactamase enzymes conferring resistance to a broad range of β-lactam antibiotics [27]. Table S3 lists eight isolates carrying *bla*_NDM_ genes from seven unique patients, with two isolates originating from the same patient (both from the same urine culture but identified as strains with slightly different AMR profiles). Given that all detected *bla*_NDM-5_ genes were carried by IncF type replicon plasmids, we analyzed pairwise SNP distances to study potential horizontal plasmid transfer (Table S4). Only plasmids from the two *E. coli* strains from the same sample fell below our predefined cutoff value (<15 SNPs/100 Kb) [28].

## Discussion

There is increasing recognition that analysis of genetic relatedness amongst bacteria causing infections in healthcare settings could substantially add to IPC efforts to mitigate pathogen acquisition [1, 18]. However, widespread use of such approaches is currently limited by a host of factors including difficulty providing timely data in a cost-effective fashion [29]. Herein, we demonstrate that a low-resource infrastructure using an ONT sequencing pipeline can produce accurate WGS information in a time frame commiserate with impactful IPC interventions.

Given its flexibility and low instrumentation requirements, ONT sequencing has long been considered as potentially impactful on infectious diseases surveillance such as its use in field-based sequencing during the 2015 Ebola outbreak [30]. However, prior to the release of the R10.4.1 flow cells and Dorado basecalling algorithm, the high ONT error rate meant that it was not sufficiently accurate to determine whether bacteria were potentially part of a transmission network [9, 20]. When analyzing a diverse array of AMR pathogens, we found that stand-alone ONT sequencing generated complete genomes that generally varied from the gold standard Illumina by only 1-2 SNPs. These data are consistent with studies emerging from other investigations using recent ONT pipelines which generally have analyzed historical cohorts [9, 20, 31]. Moreover, our data were generated prospectively and analyzed by a single operator indicating the low resource utilization of our approach. In light of the tremendous impact of HAI pathogens, modeling has indicated potential cost-savings with routine WGS [32]. However, the high costs of a core sequencing facility capable of generating Illumina data within a time frame needed for effective IPC efforts may limit its application to research centers [4, 5]. We envision that ONT sequencing could be effectively utilized by a broad variety of biomedical facilities to limit HAI transmission, effectively democratizing the integration of WGS into IPC efforts [9].

Previously, routine sequencing of a diverse array of HAI pathogens showed that particular species were more likely to meet genetic cut-offs for being potentially healthcare transmitted, which caused us to focus on a high-risk group of bacterial pathogens [4]. Still, our finding that 8.7% of such isolates met the criteria for potential transmission was lower than the 10.8% reported in the Pittsburgh, U.S.A. based study [4] or the 30% reported in a recent investigation from Australia [5]. One potential explanation is that with our relatively low number of samples, we lacked the power to fully identify clusters. Additionally, we almost exclusively analyzed clinical infection samples whereas many of the clusters identified in the Australian study came from active colonization surveillance, and thus we may have missed transmission instances that did not result in clinical disease. Finally, given the highly immunocompromised nature of our patients, robust IPC efforts at our hospital, such as routine gloves and masking when entering the rooms of certain types of patients, may mitigate transmission. In spite of some differences, a striking consistent finding between our studies and others is the very high rates of genetic relatedness amongst VREfm which was 46% in our study, 36% in the Pittsburgh study, and 92% in the Australian investigation [4, 5]. These findings are not limited to a single ST and suggest that much work remains to be done regarding limiting transmission of VREfm in healthcare settings. Conversely, unlike several recent studies, we found no clusters of closely related MRSA strains despite sequencing 80+ isolates [7, 13]. Thus, our data also suggest that WGS resources could be targeted using adaptive strategies dependent upon local findings rather than sequencing all drug-resistant pathogens.

In summary, we demonstrate that a low-resource, stand-alone ONT sequencing platform shows promise for real-time monitoring of healthcare-associated transmission and outbreak detection. Integration of such an approach into IPC efforts could assist with limiting HAIs in a wide variety of healthcare settings.

## Data Availability

All data produced in the present study are available upon reasonable request to the authors

## Acknowledgements

We thank the members of the MDACC clinical microbiology laboratory for providing isolates in timely fashion. The authors acknowledge the support of the high-performance computing research facility at the University of Texas MDACC for providing computational resources that have contributed to the research results reported in this paper. We acknowledgment some pictures were created with BioRender.com. The authors thank Andrew Yang’s help for scientific writer and editorial assistance.

## Ethical statement

This study received approval from the University of Texas MDACC Quality Improvement Assessment Board (protocol ID no. QIAB-1051).

