## Supplemental File for "Rapid Whole Genome Characterization of High-Risk Pathogens Using Long-Read Sequencing to Identify Potential Healthcare Transmission"

### Supplementary Appendix

1. Supplementary Methods
2. Figure S1: Schematic and time required for ONT sequencing workflow
3. Figure S2: Phylogenetic analysis of *E.coli* strains with sequence types
4. Figure S3: Phylogenetic analysis of *K. pneumoniae* strains with sequence types and clusters
5. Figure S4: Phylogenetic analysis of *P. aeruginosa* strains with sequence types
6. Figure S5: Phylogenetic analysis of VREfm strains with sequence types and clusters
7. Figure S6: Phylogenetic analysis of MRSA strains with sequence types and clonal complexes
8. Figure S7: Heat map of major genes mediating β-lactam resistance
9. Table S1: Summary of quality control for long-read sequencing data
10. Table S2: List of clusters detected by ONT sequencing pipeline
11. Table S3: List of plasmids carrying the $\textit{bla}_{\text{NDM}}$ gene
12. Table S4: Normalized SNP distance matrix of five $\textit{bla}_{\text{NDM-5}}$ gene-containing plasmids isolated from *E. coli*
13. Supplementary Reference

### Supplementary Methods

#### New Delhi Metallo-β-Lactamase (NDM) encoding plasmid transfer analysis

We used the assembled FASTA file from our in-house Flyest pipeline for AMRFinderPlus v3.11.14 to detect and analyze antimicrobial resistance (AMR) genes [1]. Parameters included a minimum 90% coverage of the reference protein and 98% identical amino acids in the alignment for a hit. Plasmids were identified as closed, circular contigs. Plasmid replicons were identified using staramr v0.10.0 with default parameters (https://github.com/andrewjpage/tiptoft). The annotation of horizontal plasmid transfer was conducted using Roary v3.13.0 (https://github.com/sanger-pathogens/Roary), which is based on Prokka v1.14.5 [2], and pairwise SNP distance matrix was viewed with snps-dist v0.8.2 (https://github.com/tseemann/snp-dists). The cutoff value for horizontal plasmid transfer was less than 15 SNPs per 100 kb of normalized consensus plasmid sequence [3], with the sequence length calculated from Roary's core gene alignment file, resulting in 50,457 bp in our study.

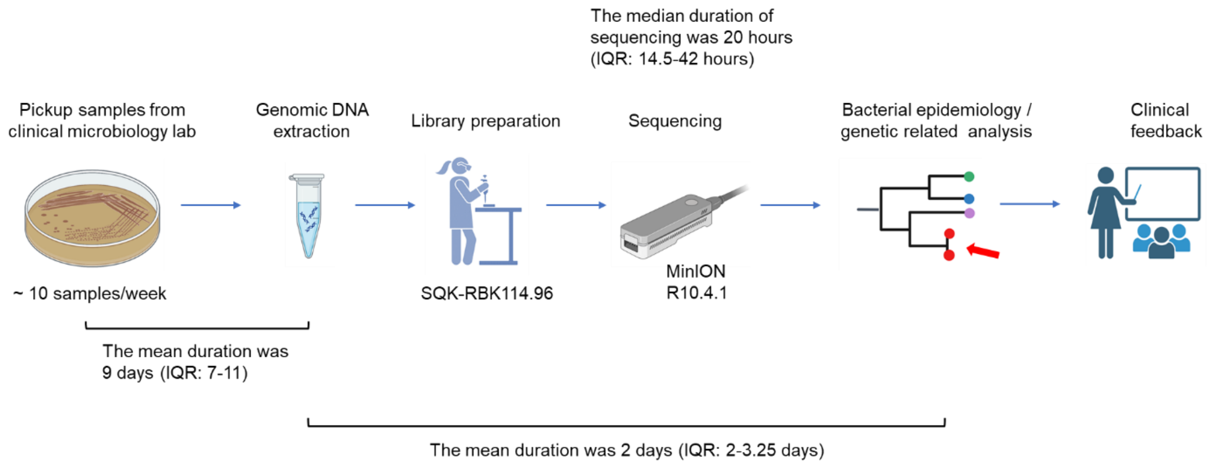


**Figure S1. Schematic and time required for ONT sequencing workflow**

Overview schematic of steps in the ONT sequencing pipeline with the mean time of each step listed in the figure.

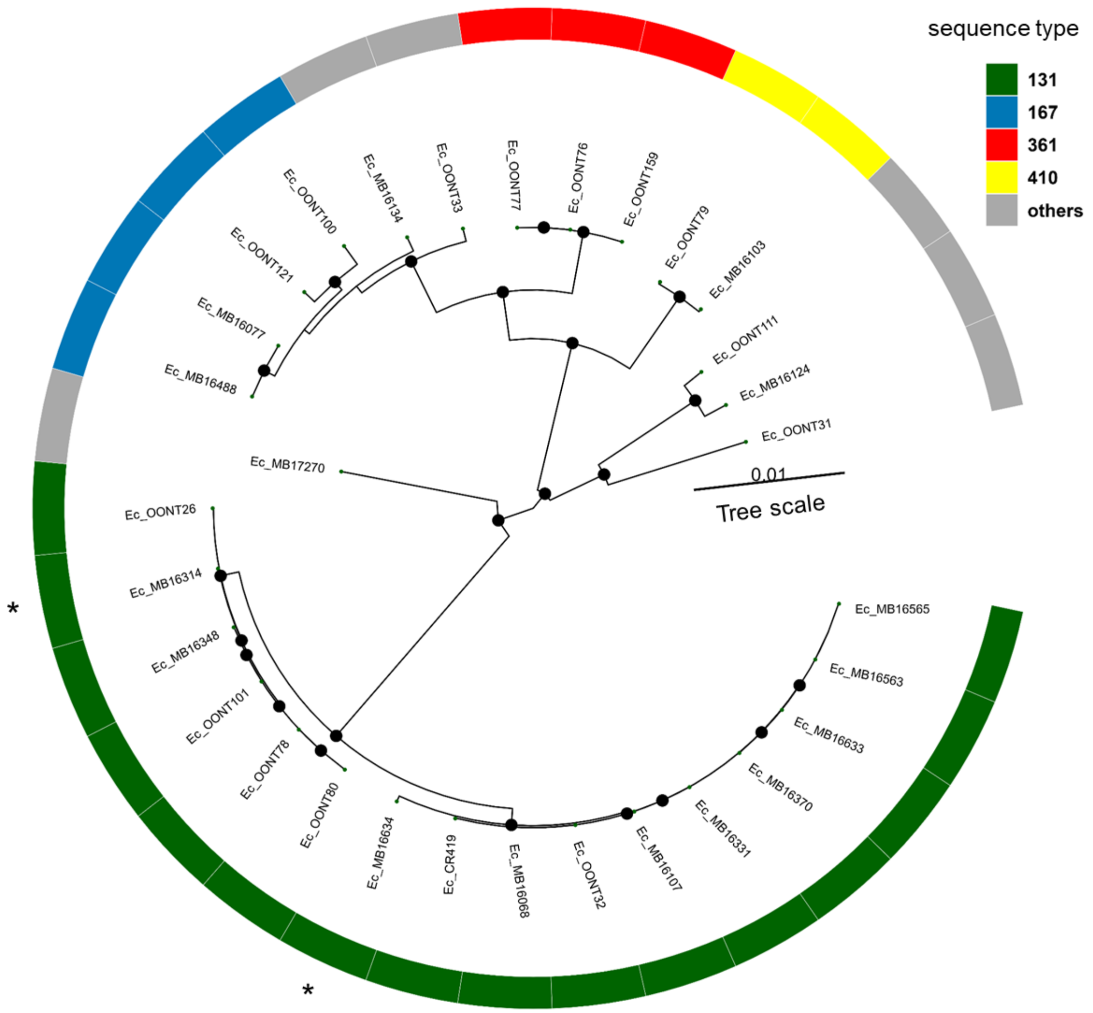


**Figure S2. Phylogenetic analysis of *E.coli* strains with sequence types**

This phylogenetic tree shows 31 *E. coli* strains, with sequence types (STs) color-coded and STs with a single locus variant marked with asterisks (*). Black circles indicate internal nodes with bootstrap values higher than 90. Genomes were annotated using Prokka v1.14.5, generating GFF3 files for subsequent pan-genome analysis. Core-gene alignments were performed with Roary v3.13.0, and a maximum-likelihood phylogeny was constructed using IQ-TREE v2.3.0 with 1000 bootstrap replicates. The resulting phylogenetic tree was visualized using the R package ggtree.

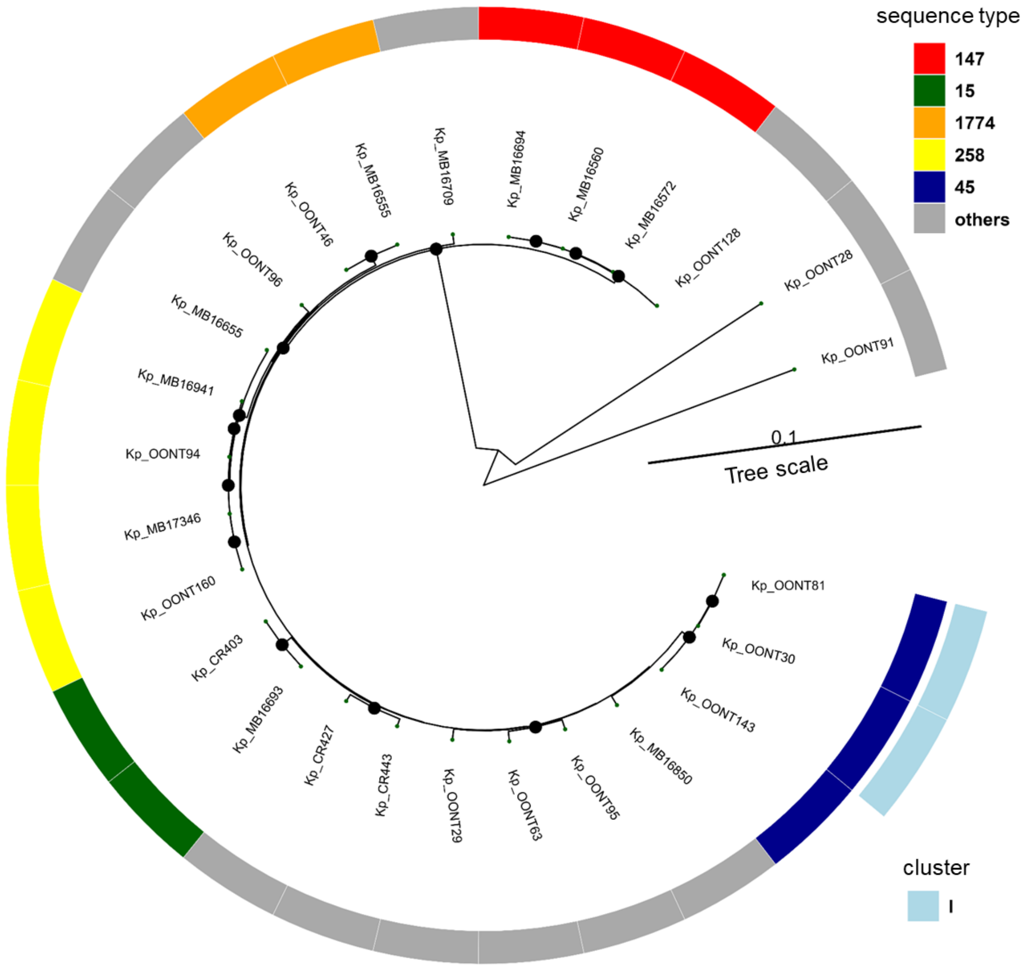


**Figure S3. Phylogenetic analysis of *K. pneumoniae* strains with sequence types and clusters**

This phylogenetic tree shows 26 *K. pneumoniae* strains, with distinct colors for different sequence types (STs) on the inner layer. The outer layer indicates cluster I, identified below the predefined cutoff value, in light blue. Black circles indicate internal nodes with bootstrap values higher than 90. Genomes were annotated using Prokka v1.14.5, generating GFF3 files for subsequent pan-genome analysis. Core-gene alignments were performed with Roary v3.13.0, and a maximum-likelihood phylogeny was constructed using IQ-TREE v2.3.0 with 1000 bootstrap replicates. The resulting phylogenetic tree was visualized using the R package ggtree.

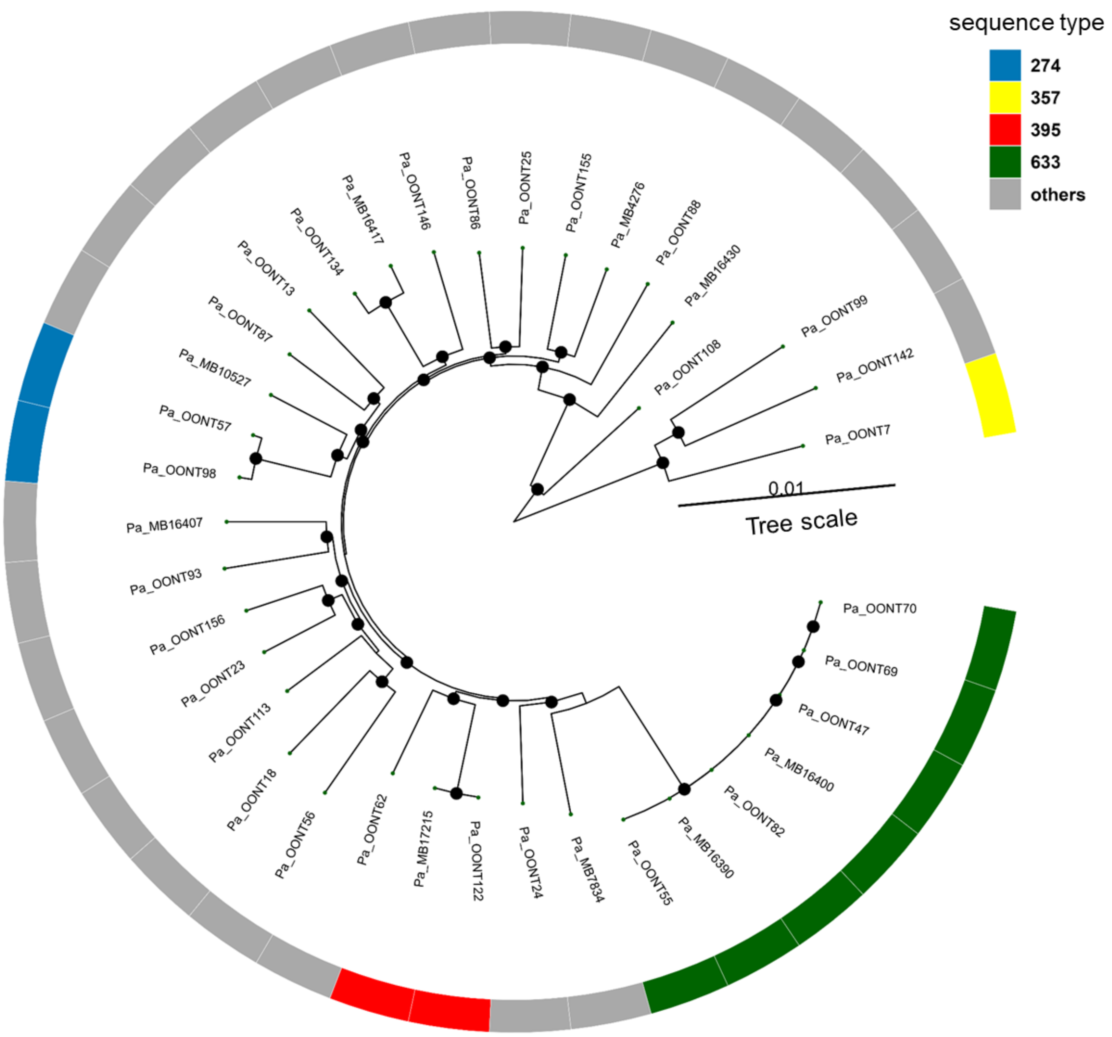


**Figure S4. Phylogenetic analysis of *P. aeruginosa* strains with sequence types**

This phylogenetic tree includes 37 *P. aeruginosa* strains, with distinct colors for different sequence types (STs). The others are all unique STs different from each other. Black circles indicate internal nodes with bootstrap values higher than 90. Genomes were annotated using Prokka v1.14.5, generating GFF3 files for subsequent pan-genome analysis. Core-gene alignments were performed with Roary v3.13.0, and a maximum-likelihood phylogeny was constructed using IQ-TREE v2.3.0 with 1000 bootstrap replicates. The resulting phylogenetic tree was visualized using the R package ggtree.

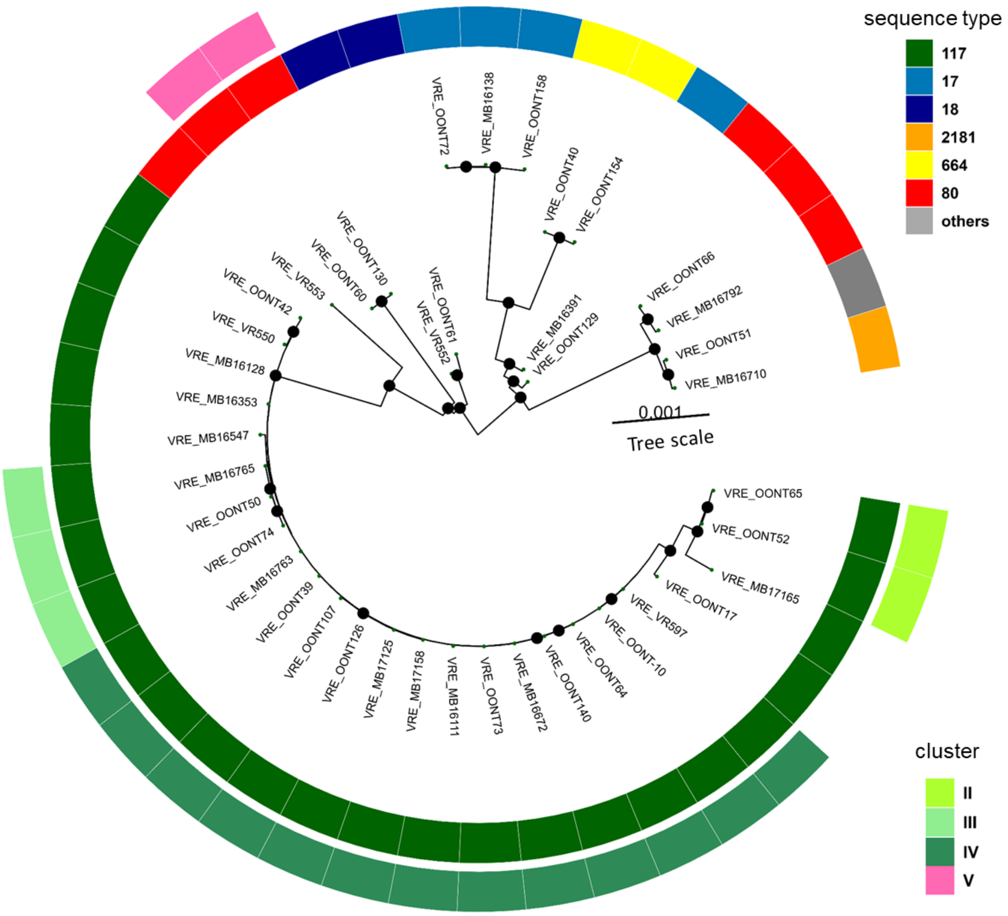


**Figure S5. Phylogenetic analysis of VREfm strains with sequence types and clusters**

This phylogenetic tree shows 41 VREfm strains, with distinct colors representing different sequence types (STs) on the inner layer. The outer layer indicates clusters identified below the predefined cutoff value, with varying shades of green for different clusters in ST117 (Clusters II, III, and IV) and pink for Cluster V, which belongs to ST80. Black circles indicate internal nodes with bootstrap values higher than 90. Genomes were annotated using Prokka v1.14.5, generating GFF3 files for subsequent pan-genome analysis. Core-gene alignments were performed with Roary v3.13.0, and a maximum-likelihood phylogeny was constructed using IQ-TREE v2.3.0 with 1000 bootstrap replicates. The resulting phylogenetic tree was visualized using the R package ggtree.

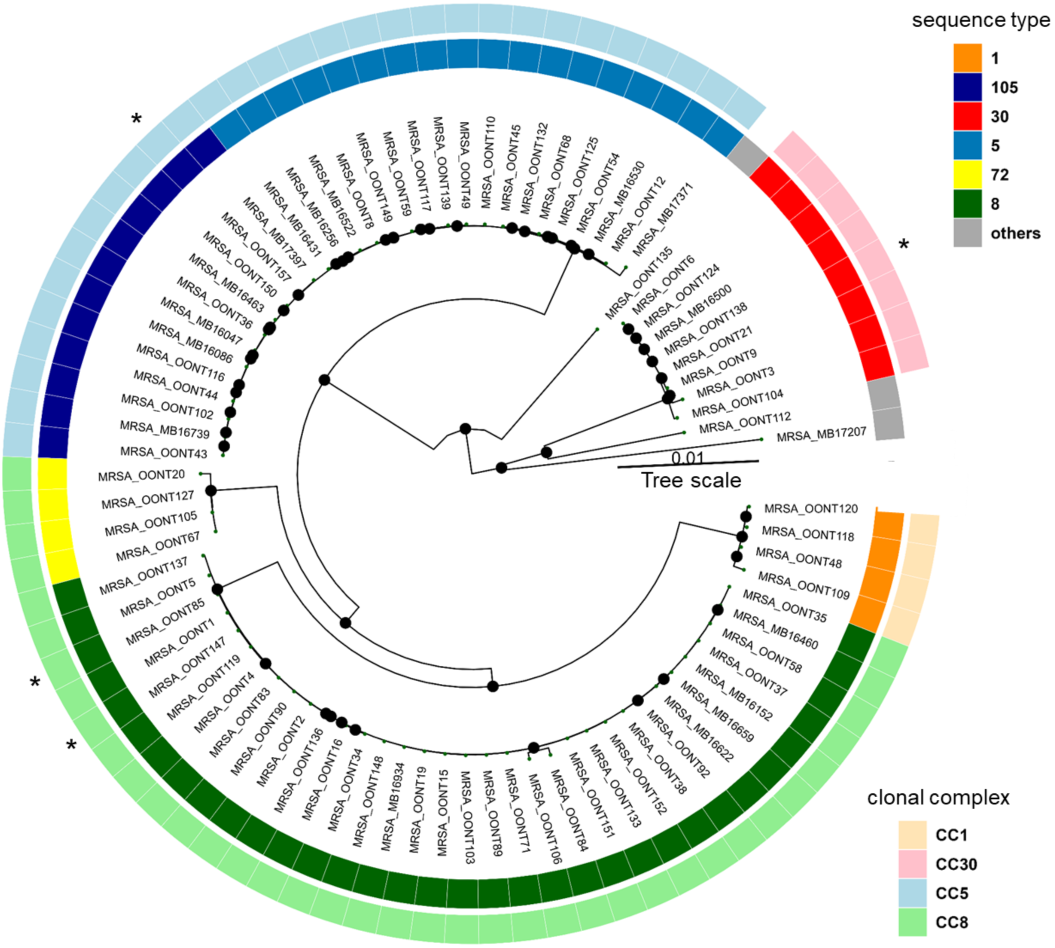


**Figure S6. Phylogenetic analysis of MRSA strains with sequence types and clonal complexes**

This phylogenetic tree includes 83 MRSA strains, with distinct colors for different sequence types (STs) on the inner layer. The outer layer indicates different clonal complexes (CCs) with varying colors. Asterisks (*) mark STs with one locus variants. Black circles indicate internal nodes with bootstrap values higher than 90. Genomes were annotated using Prokka v1.14.5, generating GFF3 files for subsequent pan-genome analysis. Core-gene alignments were performed with Roary v3.13.0, and a maximum-likelihood phylogeny was constructed using IQ-TREE v2.3.0 with 1000 bootstrap replicates. The resulting phylogenetic tree was visualized using the R package ggtree.

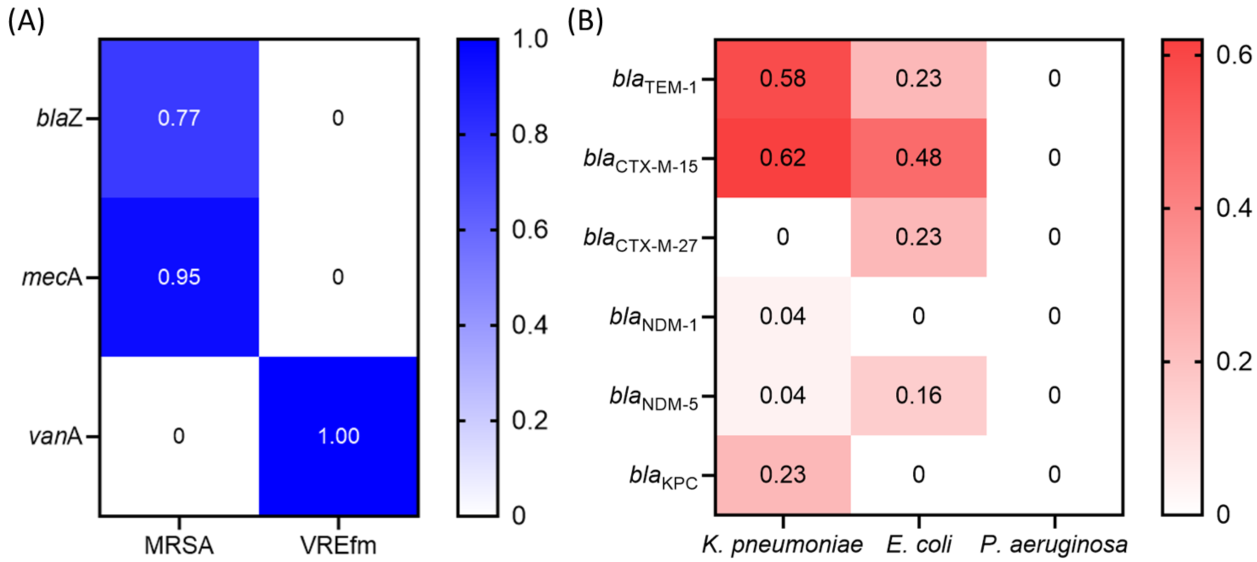


**Figure S7. Heat map of major genes mediating β-lactam resistance**

(A) Heatmap illustrating the percentage of resistance genes in MRSA and VREfm isolates. (B) Heatmap displaying the percentage of β-lactam resistance genes in *K. pneumoniae, E. coli*, and *P. aeruginosa* isolates. The color intensity represents the relative frequency of each gene, with darker shades indicating higher occurrence.

**Table S1. Summary of quality control for long-read sequencing data**

| | Total | *E. coli* | *K. pneumoniae* | *P. aeruginosa* | VREfm | MRSA | Other* |
|---|---|---|---|---|---|---|---|
| Number of isolates | 242 | 31 | 26 | 37 | 41 | 83 | 24 |
| Coverage median | 78 | 63 | 80 | 45 | 87.5 | 87 | 99 |
| (Q1, Q3) | (52, 118) | (43, 99) | (53, 125) | (38, 62) | (75, 128) | (65, 128) | (40, 135) |
| N50 (bp) | 12591 | 9419 | 12684 | 19522 | 10192 | 11571 | 13529 |
| Median ONT raw read quality (Q score) | 21.0 | 19.7 | 19.8 | 20.6 | 20.1 | 21.4 | 20.6 |
| Number of Seqsphere cgMLST target genes | | 2513 | 2358 | 3867 | 1423 | 1861 | N/A |
| Good allele target percentage average | | 99.1 | 99.1 | 99.6 | 99.0 | 98.8 | N/A |
| Good allele target percentage range | | 98.6-99.4 | 97.3-99.7 | 97-100 | 97.8-99.6 | 96.7-99.7 | N/A |

"Other*" column includes 18 *Enterobacter cloacae* complex, 3 *Acinetobacter baumannii* complex, 2 *Klebsiella aerogenes*, and 1 *Citrobacter freundii*. N/A = not applicable and refers to the fact that cgMLST schema are not currently available for all these organisms.

**Table S2. List of clusters detected by ONT sequencing pipeline**

| Organism | ST | Cluster Name | Cluster Size | Days between first and last case (days) | Average Pairwise SNPs** | Likelihood of transmission | Service |
|---|---|---|---|---|---|---|---|
| *Klebsiella pneumoniae* | 45 | I | 2 | 58 | 8 | Unlikely | NA |
| Vancomycin-resistant *E. faecium* | 117 | II | 2 | 21 | 19 | Possible | Building B |
| | 117 | III | 3 | 32 | 11 | Two patients showed probable, and one was unlikely | Ward 1 |
| | 117 | IV | 12 | 225 | 12 | Patients A, C, D, E, H, J, L* -> probable<br>Patients B, I* -> possible<br>Patients F, G, K* -> Unlikely | Ward 1<br>Ward 2<br>Ward 4 |
| | 80 | V | 2 | 80 | 5 | Possible | Ward 7 |

*The patient letter refers to Figure 4

**The number of pairwise SNPs was from MINTyper distance matrix output

**Table S3. List of plasmids carrying the $bla_{NDM}$ gene**

| Species | Sequence type | Strain | NDM-type | Sample type | The name of the plasmid carried the NDM gene | Plasmid size (bp) | Note |
|---|---|---|---|---|---|---|---|
| *Enterobacter cloacae* complex | 78 | OONT53 | NDM-1 | urine | IncM2 | 87,450 | A board-host range plasmid |
| *Klebsiella pneumoniae* | 211 | OONT95 | NDM-1 | pelvic fluid | IncHI2/IncHI2A | 311,912 | Hybrid super-plasmid carried both $bla_{NDM-1}$ and *mcr-9* gene |
| *Klebsiella pneumoniae* | 11 | MB16655 | NDM-5 | blood | IncFIB(pNDM-Mar) /IncHI1B(pNDM-MAR) | 375,444 | Hybrid plasmid encoding antimicrobial resistance and virulence genes |
| *E. coli* | 361 | OONT76 | NDM-5 | urine | IncFII | 103,609 | From the same patient and sample |
| *E. coli* | 361 | OONT77 | NDM-5 | urine | IncFII | 98,137 | |
| *E. coli* | 167 | OONT100 | NDM-5 | urine | IncFIA/IncFIB | 140,363 | potential novel IncFIA/IncFIB hybrid plasmid with $bla_{NDM-5}$ and $bla_{CTX-M-15}$ |
| *E. coli* | 167 | OONT121 | NDM-5 | blood | IncFIA | 125,822 | plasmid with $bla_{NDM-5}$ and $bla_{TEM-1}$ |
| *E. coli* | 361 | OONT159 | NDM-5 | blood | IncFIA | 157,711 | plasmid with $bla_{NDM-5}$ and $bla_{CTX-M-15}$ |

**Table S4. Normalized SNP distance matrix of five *bla*$_{NDM-5}$ gene-containing plasmids isolated from *E. coli***

| | Ec_OONT100 IncFIA/IncFIB | Ec_OONT121 IncFIA | Ec_OONT159 IncFIA | Ec_OONT76 IncFII | Ec_OONT77 IncFII |
|---|---|---|---|---|---|
| Ec_OONT100 IncFIA/IncFIB | 0 | 583 | 202 | 452 | 464 |
| Ec_OONT121 IncFIA | 583 | 0 | 616 | 593 | 605 |
| Ec_OONT159 IncFIA | 202 | 616 | 0 | 523 | 535 |
| Ec_OONT76 IncFII | 452 | 593 | 523 | 0 | 12 |
| Ec_OONT77 IncFII | 464 | 605 | 535 | 12 | 0 |
